# Bedside three-dimensional acoustic angiography and perfusion imaging for early detection of delayed cerebral ischemia in aneurysmal subarachnoid hemorrhage: A Feasibility study

**DOI:** 10.1101/2025.05.18.25327760

**Authors:** Clément Gakuba, Louise Denis, Georges Chabouh, Maxime Gauberti, Jean-Denis Moyer, Tom Gavet, Joséphine Malczuk, Nathalie Laquay, Sylvain Bodard, Arthur Chavignon, Vincent Hingot, Denis Vivien, Olivier Couture

## Abstract

Aneurysmal subarachnoid hemorrhage (aSAH) is a life-threatening condition with high rates of secondary complications such as delayed cerebral ischemia (DCI). Early detection of perfusion deficits is critical but remains challenging with conventional imaging modalities that are intermittent, non-portable, and provide limited temporal resolution. Here, we evaluate a novel investigational platform for volumetric acoustic angiography and bedside cerebral perfusion imaging using three-dimensional contrast-enhanced ultrasound (3D CEUS). In a prospective feasibility study, eleven aSAH patients underwent bilateral 3D CEUS at admission, day 5, and day 10 in the neurocritical care unit. The system enabled reconstruction of 3D microvascular volumes and time-intensity evolution (TIE) curves for cerebral blood flow (CBF) assessment. Global perfusion measurements were successful in 95% of acquisitions. Notably, TIE-derived perfusion patterns at day 5 showed strong correlation with poor neurological outcome (88.9% accuracy) suggesting the prognostic potential of the technique. This study demonstrates, for the first time, the feasibility of bedside volumetric CEUS for dynamic assessment of cerebral perfusion in patients with aSAH. The method offers high spatial and temporal resolution, real-time feedback, and correlation with clinical trajectory. These findings position three dimensional CEUS-based acoustic angiography as a promising tool for early detection of DCI and improved neuromonitoring in the intensive care unit.

## Introduction

Aneurysmal subarachnoid hemorrhage (aSAH) accounts for approximately 5% of all strokes, with an incidence of about 10 cases per 100,000 people annually^1^. Despite its relatively low prevalence, aSAH is responsible for over a quarter of stroke-related life years lost before age 65, imposing a significant economic and societal burden. Mortality rates can reach 60% in some cohorts, and more than half of survivors experience persistent disabilities or cognitive deficits, significantly impacting their quality of life. For decades, it has been recognized that some patients who survive the initial hemorrhage later deteriorate neurologically, leading to either death or long-term deficits. The development of radiological imaging in the 1950s revealed the occurrence of vasospasm following the initial hemorrhage, which was quickly suspected as the main cause of secondary neurological decline^2^. This led to the investigation of vasodilator therapies, culminating in the approval of nimodipine to reduce delayed neurological deficits. However, 40 years later, no effective therapeutic strategy has emerged to significantly improve outcomes in aSAH patients^3^. In patients suffering from SAH, about half will experience some form of delayed hypoperfusion, worsening outcomes in 20–30% of cases^4^. Delayed cerebral ischemia (DCI) typically occurs around 7 days post-hemorrhage, with peak risk on the fifth day^5^. While cerebral vasospasm is frequently implicated, secondary neurological decline is increasingly recognized as a multifactorial process involving hypoperfusion, microcirculatory dysfunction, and metabolic failure. Broadly, aSAH patients fall into two groups: those with assessable clinical evaluations and those without, with secondary neurological deficits being more frequent and severe in the latter. In the first group, deficits are identified through clinical examination, guiding treatment strategies such as induced arterial hypertension and endovascular interventions^6^,^7^. In the second group, Transcranial Doppler (TCD) has been a standard bedside tool since the 1980s, used to infer cerebral blood flow based on velocity measurements^8^,^9^. However, TCD does not provide direct information on vessel diameter or blood flow, making it an imperfect surrogate for cerebral perfusion. Since Doppler velocity depends on the ratio of blood flow to vessel diameter, its interpretation relies on assumptions about vessel caliber, which are often flawed^10^.

Advanced neuroimaging techniques such as Computed Tomography Angiography (CTA)^11^ and CT Perfusion (CTP)^12^,^13^ provide detailed insights into cerebral perfusion and vascular status. However, their utility is limited by logistical challenges, including restricted access, radiation exposure, and the impracticality of frequent bedside use. As a result, the search for more effective neuromonitoring tools continues. Existing modalities tend to fall into two categories: those that detect the downstream effects of ischemia, such as EEG or intracranial pressure monitoring, which are specific but often lack sensitivity; and those that target arterial narrowing, such as Transcranial Doppler (TCD) or CTA, which are sensitive but not always specific. Although the role of CTP in guiding clinical decisions remains under discussion, there is broad agreement that monitoring local cerebral blood flow is key to enabling timely and targeted treatment^14–21^. Yet, the limited repeatability and poor accessibility of CTP render it far from ideal in dynamic, acute care settings.

To overcome the limitations of traditional imaging, ultrasound presents an attractive alternative: it is portable, widely accessible, and compatible with bedside use—key features for dynamic neuromonitoring in critical care^22^. Contrast Enhanced Ultrasound (CEUS), which relies on intravenously injected microbubble contrast agents, is already well-established in clinical practice for abdominal applications, particularly in the liver and kidneys^23^,^24^. From CEUS acquisitions, tissue perfusion can be assessed by tracking the spatiotemporal distribution of contrast—an approach known as Ultrasound Perfusion Imaging (UPI). UPI could serve as a point-of-care alternative to Computed Tomography Perfusion (CTP). Similar to CTP, UPI quantifies cerebral blood flow by analyzing Time-Intensity Evolution (TIE) curves as contrast passes through brain tissue^25^,^26^. It has shown promise in acute ischemic stroke^27–29^, pediatric neuromonitoring^30^, and vasospasm following SAH^31^. Despite encouraging results, its clinical adoption remains experimental, likely due to the absence of dedicated commercial devices. However, 2D CEUS suffers from intrinsic shortcomings: high user dependency, subjective plane selection, and sensitivity to out-of-plane motion and flow, all of which reduce reproducibility and complicate longitudinal monitoring. Additionally, long acquisition times required for reliable perfusion quantification can be difficult to standardize in a clinical setting^32^. In this study, we address these challenges by evaluating a novel investigational system for 3D CEUS brain imaging. This device enables the reconstruction of 3D Microvascular Imaging (MVI) volumes (Fig1A), offering both angiographic visualization and quantitative perfusion analysis through TIE curves (Fig 1B). We performed a prospective feasibility study in 11 patients with aSAH, assessing its ability to capture cerebral vascular morphology and extract clinically relevant perfusion metrics for bedside neuromonitoring (Fig1C).

**Figure 1.**
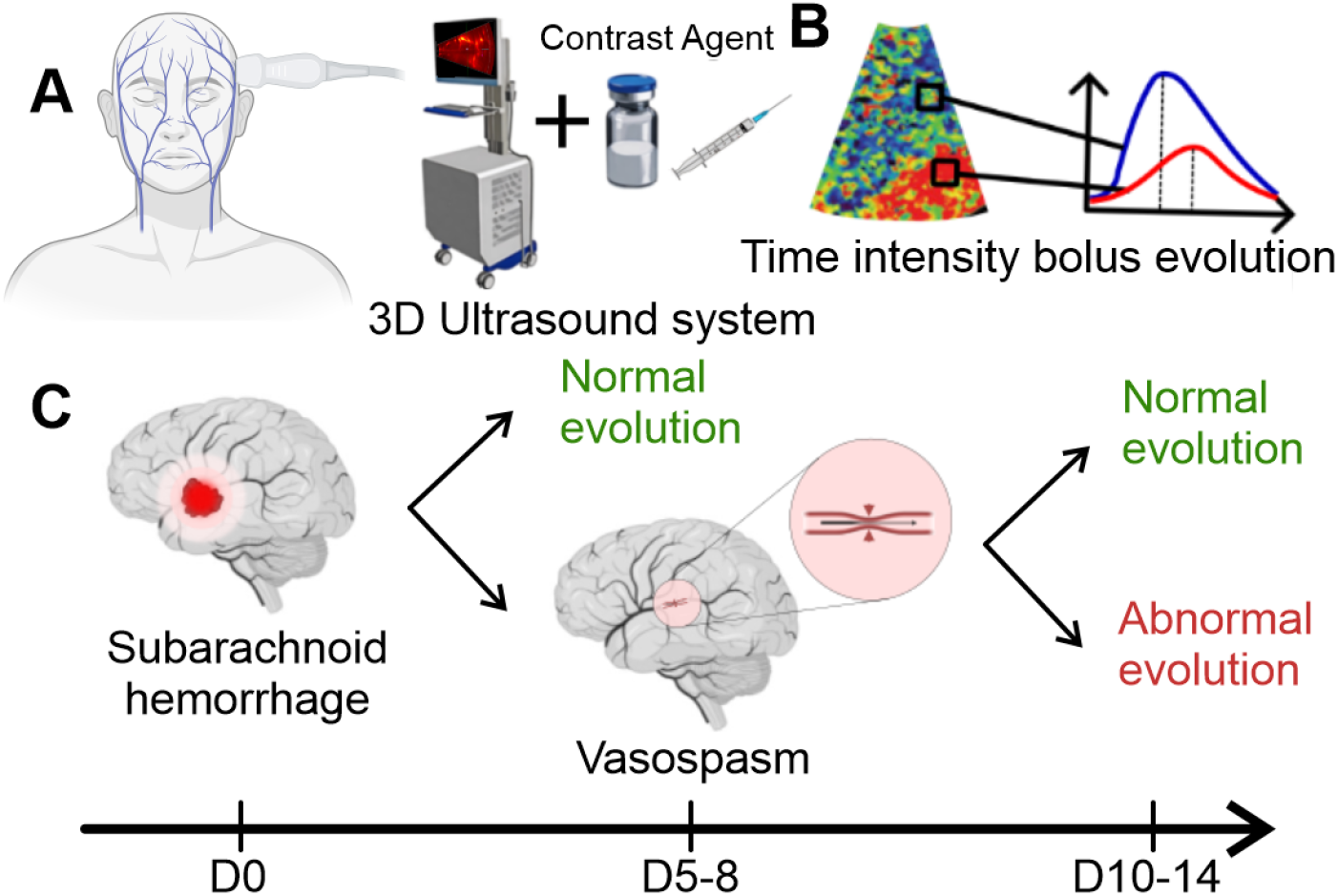
Schematic representation of 3D transcranial contrast-enhanced ultrasound (CEUS) monitoring in patients with subarachnoid hemorrhage (SAH): (A) A 3D ultrasound system is used in combination with a bolus injection of contrast agent to enhance vascular imaging. (B) Time intensity evolution monitors the bolus and enables quantitative assessment of cerebral perfusion in 3D. (C) The proposed monitoring protocol follows SAH progression from the acute phase (D0) to the vasospasm phase (D5–8) and late-stage evolution (D10–14). Depending on perfusion dynamics, patients may show either normal evolution (green) or abnormal evolution (red), indicating worsening vascular conditions.

## Results

### Patient characteristics and clinical study design

The study design included 3 imaging sessions where two 3D acquisitions are performed through the left and right temporal windows. The first session is planned in the first 72h following the onset of the SAH after the initial treatment of the aneurism. The second session is performed on the fifth day, where the risk of vasospasm is the highest. The third session is performed between days ten and fourteen after the high-risk phase.

Ninety-five (95%) of acquisitions (63/66) exhibited contrast enhancement following the injection of contrast, i.e. the perfusion curve has increased by at least 15% compared with the baseline intensity. The acquisitions where the enhancement could not be observed were explained respectively by a lack of a temporal window, inadequate probe positioning and known absence of perfusion. No adverse effects were observed during the study. Acquisitions were performed by positioning the probe on the patient’s head, just anterior to the temporal window. Continuous diverging waves were used to track the passage of the bolus injection of microbubble contrast agents spanning 9×8×8 *cm*^3^ at 1000 volumes/sec. Volumetric images were registered with those acquired via CT and MRI as part of the patient’s management.

### Acoustic angiography with the Micro Vessel Imaging (MVI) reconstruction

Based on the MVI reconstruction, the main cerebral arteries were identifiable in most patients. From the temporal window, the field of view contains the circle of Willis as well as the Internal Carotid Artery (ICA), the proximal section of the Anterior Cerebral Artery (ACA) and of the Posterior Cerebral Artery (PCA) and most of the Middle Cerebral Artery (MCA), even some more distal segments. The variability in detectable structures was mainly influenced by probe positioning and the patient’s condition.

Fig.2 illustrates representative examples of 3D acoustic angiography acquisitions, highlighting variability in cerebral artery detection. Figure 2A shows a complete vascular reconstruction with clear identification of all major arteries, including ICA, ACA, basilar artery, MCA (M1 and M2 segments), and PCA, indicating an optimal acquisition. Figure 2B presents an example of posterior probe placement, where only vessels from the posterior circulation are captured. Several slices of the 3D CEUS are shown in Fig.Supplementary 1 and Supplementary Video 1. For anatomical reference, the schematic in Figure 2C shows the Circle of Willis and corresponding labeled arteries. In a subset of acquisitions, registration with angiographic CT allowed for cross-modality comparison; Figure 2D demonstrates strong visual concordance between the acoustic angiogram and the CT scan. Quantification of arterial detection across all subjects is shown in Figure 2E, confirming consistently high detection rates for the ICA and MCA1 segments, with lower rates for more distal branches and the ACA.

**Figure 2.**
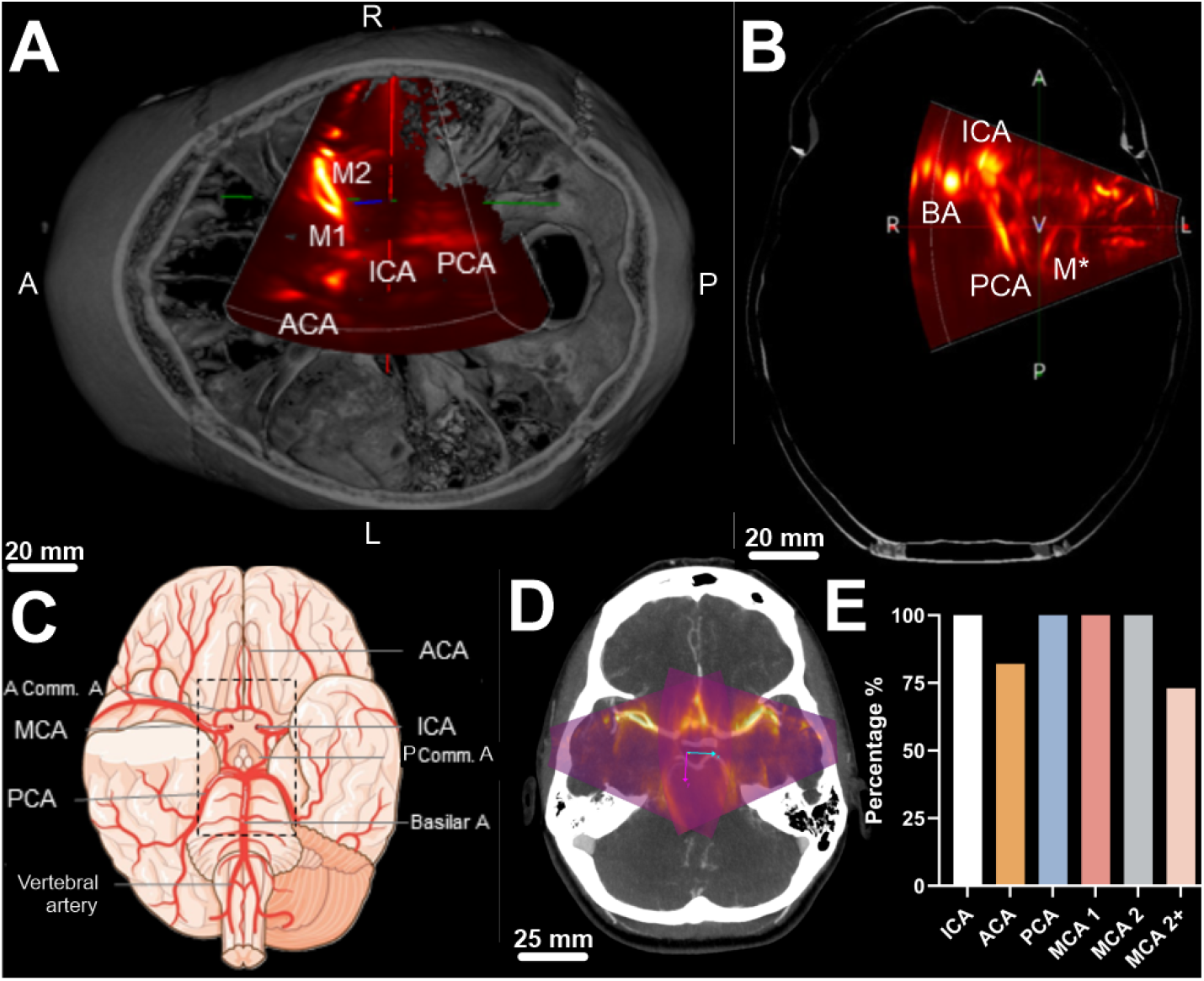
Identification and visualization of cerebral vasculature using acoustic angiography with Microvascular Imaging (MVI). (A–B) Axial views of a 3D MVI overlaid on anatomical images showing the localization of major cerebral arteries, including the anterior cerebral artery (ACA), middle cerebral artery (MCA; M1 and M2, M* segments), internal carotid artery (ICA), Basilar Artery (BA) and posterior cerebral artery (PCA). (C) Schematic view of the human brain vasculature for anatomical reference. (D) Overlay of the MVI on an angio-CT slice, highlighting transcranial imaging capabilities and deep vessel detection. (E) Vessel detection rates for major cerebral arteries across all imaging sessions (n = 33), showing high consistency in identifying the ICA, ACA, PCA, and MCA segments (M1 and M2).

### Perfusion imaging and analysis of the TIE curves

Figure 3 shows two examples of perfusion imaging in a patient with no clinical sign of cognitive deficit, referred to as “normal” perfusion (Fig 3.A-i,ii,B and C), and a patient with clinical signs of cognitive deficit (here following a confirmed vasospasm), referred to as “abnormal” perfusion (Fig 3.D-i,ii,E and F). The perfusion is characterized as maps of Time To Peak (TTP) and Mean Transit Time (MTT), but also histograms showing the spread in TTP and MTT values within the images. In the “normal” perfusion, the values are more homogeneous for TTP (Fig 3.A-i,ii) and MTT (Fig 3.B) that for the “abnormal” perfusion (Fig 3.D-i,ii & Fig 3.E). The heterogeneity can be seen on the images, and a typical difference of TTP and MTT of several seconds can be observed within just a few centimeters of brain tissue.

**Figure 3.**
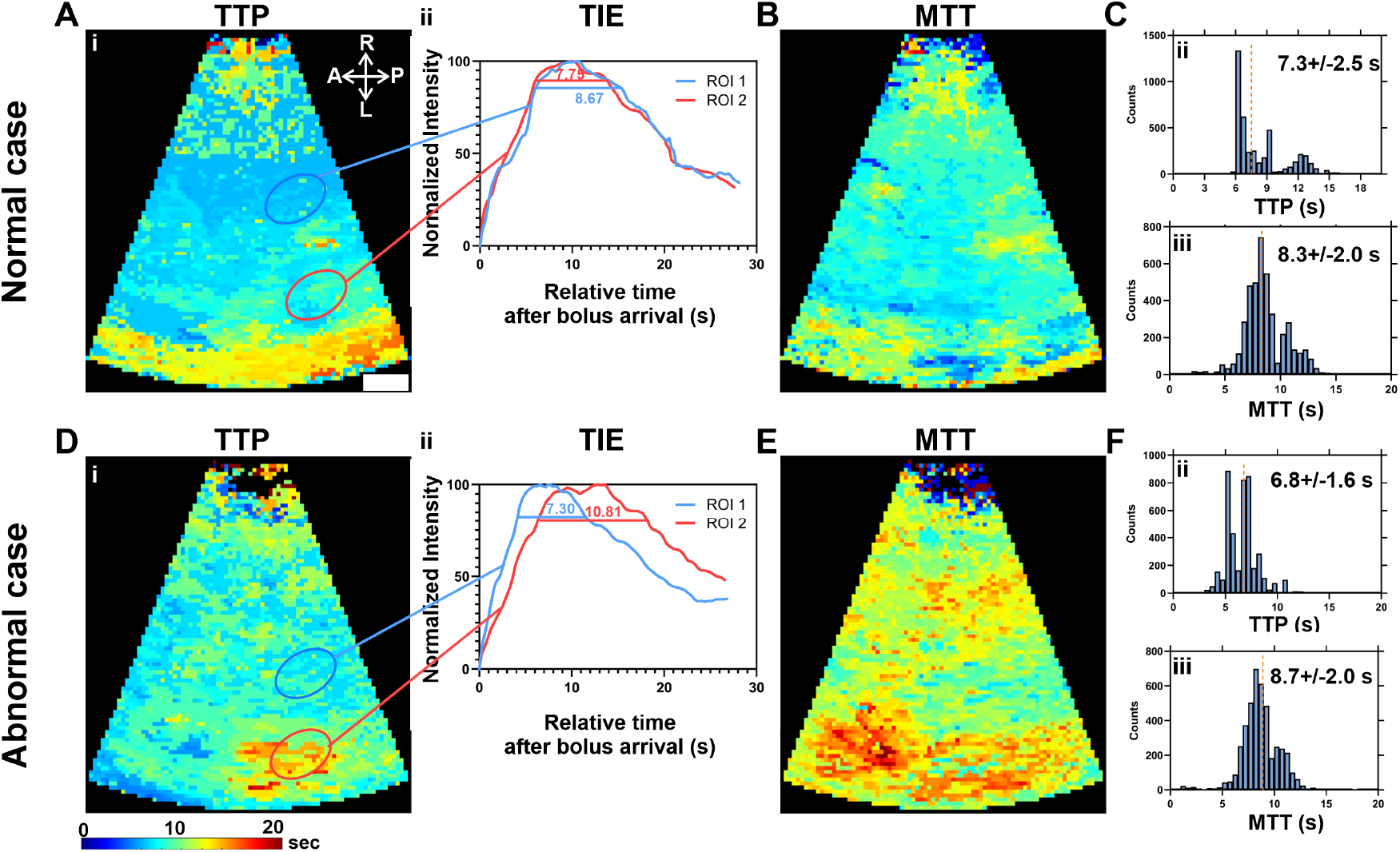
Evaluation of cerebral perfusion using Time-to-Peak (TTP) and Mean Transit Time (MTT) maps derived from 3D CEUS in different vascular territories. (A-B) TTP and MTT maps, respectively, from a case with normal perfusion. Color-coded parametric maps (i) represent perfusion delays in seconds, with blue indicating early enhancement and red reflecting delayed flow. Corresponding time intensity evolution (TIE) curves (ii) from selected ROIs (circled in blue and red) show overlapping and symmetric wash-in profiles, confirming normal hemodynamics. The histograms C-ii,iii illustrate the TTP and MTT, with median times of 7.3 *±* 2.5 s and 8.3*±* 2.0 s, respectively. Panel D and E present TTP and MTT maps, respectively, from a case with abnormal perfusion. The TIE curves (ii) show a marked delay in one ROI, consistent with delayed perfusion, and the histograms F-ii,iii illustrate the TTP and MTT, with median times of 6.8 *±* 1.6 s and 8.7 *±* 2.0 s, respectively. ROIs were carefully selected to avoid large vessels, focusing instead on parenchymal regions to accurately reflect microvascular perfusion dynamics. All TIE curves are time-aligned, with 0 seconds corresponding to the beginning of the contrast bolus rise (i.e., contrast arrival), allowing for standardized comparison across patients. The white scale bar indicates 25 mm.

### Monitoring the occurrence of DCI following SAH

Figure 4 illustrates a case study of favorable neurological evolution following aSAH, where both TTP (Fig.4A) and MTT (Fig.4B) maps show stable, symmetric trends across the left and right hemispheres at three distinct time points: ICU admission, day 5, and 10 post-hemorrhage. The perfusion parameters remain consistent across both hemispheres, with no significant asymmetry or deviations. TIE curves (Fig.4C), normalized over time for each hemisphere, highlight preserved perfusion dynamics (single peak), reflecting stable cerebral blood flow. This case demonstrates a favorable clinical course with minimal changes in perfusion parameters, consistent with a patient showing good neurological progression.

**Figure 4.**
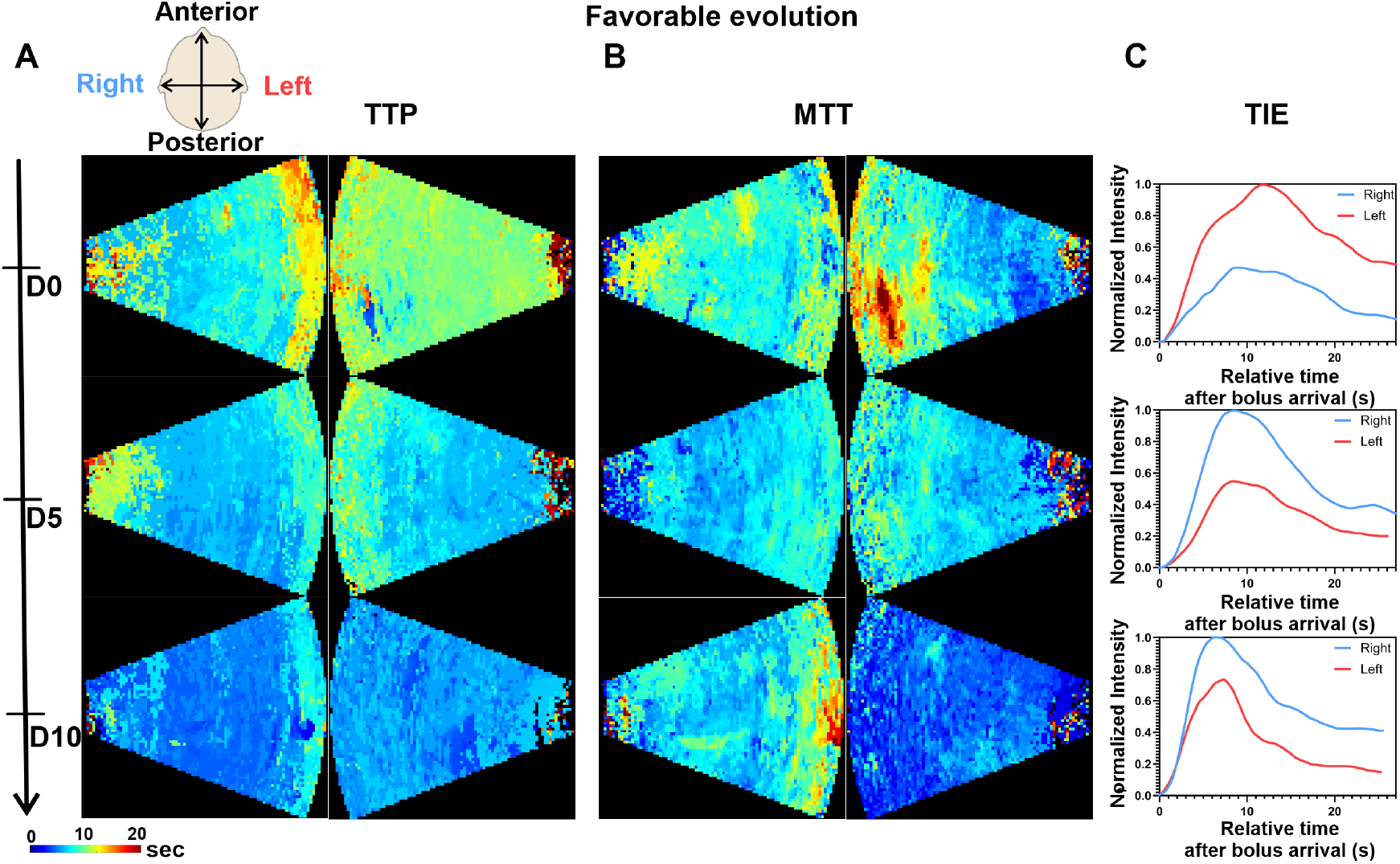
Case study: Favorable neurological evolution following aSAH. A TTP maps and B MTT maps for the left and right hemispheres at three time points: upon ICU admission, at day 5 post-hemorrhage and at day 10. C Corresponding to the global TIE over time for each hemisphere at the same time points, illustrating preserved and symmetric perfusion dynamics throughout the longitudinal study period.

Figure 5 presents a case study with unfavorable neurological evolution following aSAH. At the three time points—ICU admission, day 5, and day 10 post-hemorrhage—both TTP (Fig.5A) and MTT (Fig.5B) maps exhibit increasingly disrupted perfusion patterns (delayed bolus peak). By the final time point, the maps are predominantly characterized by noise, which reflects the patient’s clinical deterioration. This noise is indicative of poor perfusion, which aligns with the patient’s worsening condition and eventual death. The progressive deterioration in perfusion signals contrasts sharply with the earlier stable perfusion patterns observed during the initial post-hemorrhage period, underscoring the correlation between perfusion disruption and clinical decline (Fig.5C).

**Figure 5.**
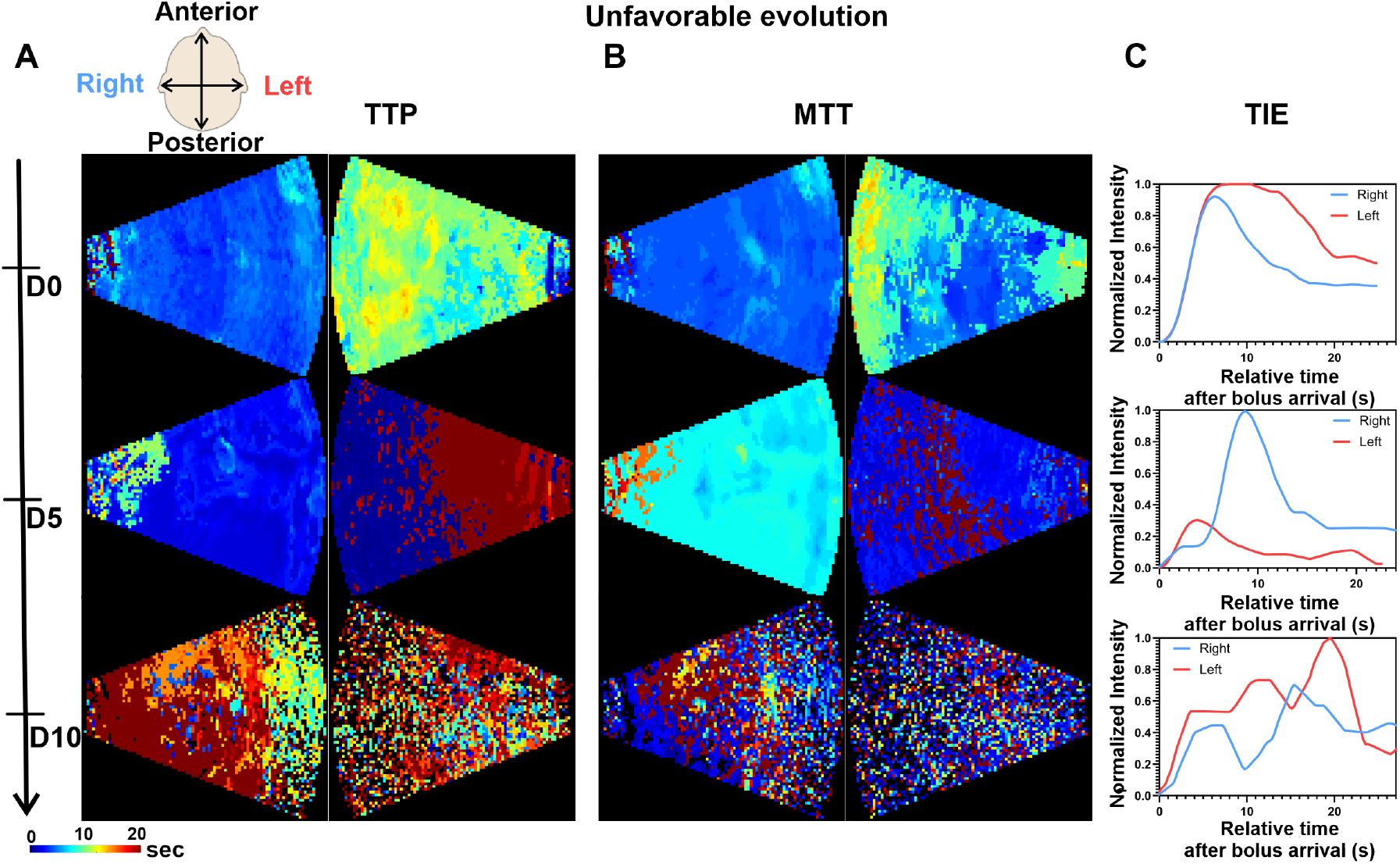
Case study: Unfavorable evolution following aSAH. A TTP and B MTT maps for the left and right hemispheres at three time points: upon ICU admission, at day 5 post-hemorrhage, and day 10. C Corresponding to the global TIE over time for each hemisphere. The final time point is characterized by high noise levels, reflecting the patient’s deterioration prior to death hence no ultrasound image is shown.

Figure 6 shows the relationship between mid-term perfusion dynamics and neurological outcomes. In Fig.6A TIE curves at time point 2 illustrate the variability in cerebral perfusion profiles across patients. These curves capture differences in the timing and amplitude of contrast enhancement, suggesting potential early markers of clinical trajectory. Fig.6B presents a Sankey diagram linking volumetric UPI interpretations to clinical outcomes. Among the 9 patients with a defined UPI classification (Favorable or Unfavorable), 88.9% showed agreement between perfusion profile and neurological outcome. Favorable UPI profiles were associated with favorable outcomes in all cases (3/3), while 83.3% of unfavorable profiles (5/6) corresponded to unfavorable outcomes. The remaining 2 patients had inconclusive UPI profiles, both of whom had favorable outcomes. These results support the potential prognostic value of volumetric UPI at mid-term timepoints. In contrast, standard TCD evaluation as recommended in the 2023 AHA guidelines for SAH^33^ — defines vasospasm suspicion at flow velocities exceeding 200 cm/s, yet demonstrates poor predictive performance in our cohort (see Supplementary Table 1).

**Figure 6.**
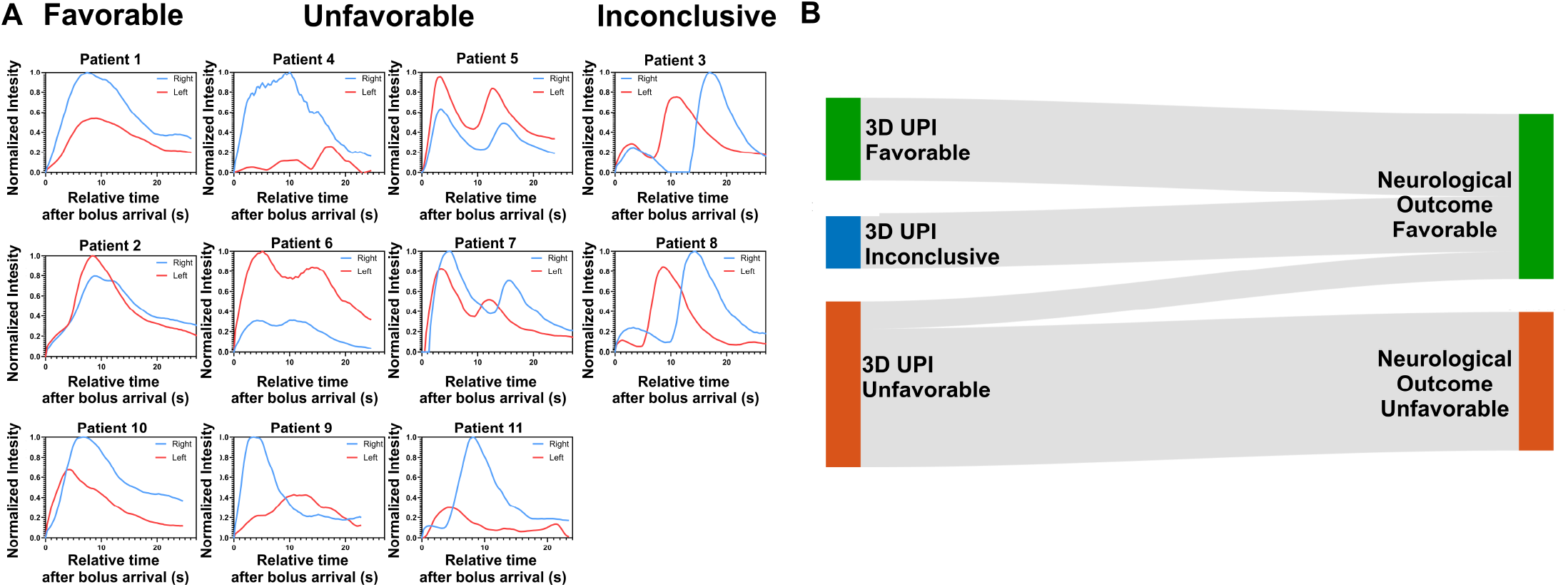
Longitudinal evolution of all patients and correlation with neurological outcomes. A TIE curves for all patients at time point 2 (Day 5), illustrating inter-patient variability in perfusion dynamics. Each curve represents one hemisphere, capturing both the timing and amplitude of contrast arrival and washout. B Sankey diagram showing the relationship between perfusion profile at time point 2 and the corresponding neurological outcome at discharge. The flow illustrates how early perfusion patterns align with favorable or unfavorable clinical trajectories.

## DISCUSSION

This study evaluated the feasibility and clinical relevance of bedside volumetric CEUS for dynamic cerebral perfusion imaging in patients with aSAH. Using a high volume-rate ultrasound system with contrast agents, we conducted a prospective study in the ICU. Our findings demonstrate that this approach is safe, repeatable, and capable of capturing both structural and functional perfusion information relevant to patient monitoring. Perfusion analysis revealed clinically meaningful differences between patients. Figure 3 contrasted two representative cases: one with normal neurological status and homogeneous perfusion, and another with cognitive deficits and heterogeneous, delayed perfusion. The corresponding TTP and MTT histograms highlighted spatial variability in cerebral blood flow, which was markedly more pronounced in the symptomatic patient. These results show that CEUS can detect perfusion disturbances, including distal branches beyond the MCA’s first segment. Longitudinal monitoring confirmed the method’s ability to track perfusion over time (Fig.Supplementary 2). In Figure 4, a patient with favorable outcome displayed stable and symmetric perfusion patterns across hemispheres over three time points (ICU admission, day 5, day 10). The corresponding TIE curves showed preserved perfusion dynamics with consistent peak timing and amplitude. In contrast, Figure 5 illustrated a case of clinical deterioration, where perfusion maps became increasingly noisy and uninformative, consistent with the patient’s worsening condition and eventual death. This progression highlights the sensitivity of CEUS to dynamic changes in cerebral perfusion. Perfusion patterns at day 5 were predictive of neurological outcomes. Figure 6 demonstrated that mid-term TIE profiles could stratify patients according to clinical trajectory. In 88.9% of cases, UPI classifications were concordant with 6-month neurological outcomes. All patients with favorable UPI profiles achieved good outcomes, while most with unfavorable profiles experienced poor recovery. These findings suggest that CEUS-derived perfusion dynamics may serve as early biomarkers of neurological prognosis following aSAH. The technique integrates well into neurocritical care workflows. Exams are well-tolerated, require only two operators, and can be completed in under 30 minutes at the bedside, avoiding the risks of patient transport. The combination of affordability, portability, and added functional insight makes CEUS a practical alternative to more expensive and less accessible modalities like CTP. This study lays the groundwork for further development and validation of volumetric UPI as a neuromonitoring tool. Future research should focus on optimizing the technique, expanding the sample size, and conducting comparative studies with established imaging modalities. The ultimate goal is to integrate UPI into standard clinical practice, providing a non-invasive, bedside solution for monitoring cerebral blood flow in SAH patients. Several limitations were identified in the study. The positioning of the probe varied between acquisitions, and although the probe’s field of view is relatively large, it was not standardized. Motion artifacts were observed in a few acquisitions but did not compromise image readability. In some acquisitions, a temporal widening of the TIE in arterial regions was observed, which was not linked to perfusion deficits. This was likely due to the slow injection of the bolus or intravenous spreading, which is typical in perfusion CT and needs to be standardized. To further understand the correlation between the perfusion data obtained with ultrasound and CT or MRI, it is important to consider the underlying hypotheses and calculation methods. Microbubbles, unlike iodine or gadolinium, are physical agents that are much larger and fewer in number. This raises issues of spatial sampling and detectability in the smallest vessels. However, in this study, we observed homogeneous contrast uptake in the parenchyma, suggesting that this may not be a significant issue. Another limitation is the calculation method. Unlike absorption in CT, the microbubble signal needs to be extracted from surrounding tissues, limiting the confidence in the signal intensity. Therefore, only temporal information obtained from the curves is usually used. In particular, transit time is typically calculated as the Full Width Half Maximum of the curve. In this study, we found that the choice of threshold can greatly impact the reading, especially with altered transits and recirculation. Therefore, we chose 80%, but this fails to describe the very slow transits situations where the pic is sometimes not even observed. In most pathologies, particularly in SAH, the heterogeneity of cerebral hypoperfusion is a critical parameter for understanding the physiology and prognosis of patient outcomes. However, the intrinsic variability of pixel-wise TIE curves makes it challenging to precisely estimate spatial and temporal resolution. The technique appears capable of imaging large perfusion deficits with temporal differences of several seconds over a few cubic centimeters. However, further studies are needed to quantify the precision.

In conclusion, this study demonstrates the feasibility and potential of 3D contrast-enhanced acoustic angiography and perfusion imaging as a bedside neuromonitoring tool for SAH patients. The safety, technical feasibility, and preliminary clinical correlations are encouraging, and further research is warranted to fully realize the benefits of this innovative approach. However, the sample size was relatively small, which limits the generalizability of the findings. Future studies with larger cohorts and refined protocols are necessary to validate these initial results.

## MATERIALS AND METHODS

### Patient characteristics and clinical study design

This study was a phase 1 clinical trial (NCT06793839). Nineteen patients diagnosed with subarachnoid hemorrhage (SAH) were screened for inclusion. Of these, 18 had aneurysmal SAH and one had non-aneurysmal SAH. The inclusion criteria focused on patients with anterior circulation aneurysms and symptom onset within 72 hours. Thirteen patients completed the full study protocol, while two did not complete all three ultrasound imaging sessions.

### Clinical management

All SAH patients were treated according to a local standard protocol in accordance with published recommendations. Initial diagnosis of aneurysmal SAH was made using non-contrast computed tomography (CT), lumbar puncture (LP) or magnetic resonance imaging (MRI). Once aneurysmal SAH was confirmed, CT angiography (CTA) or digital subtraction angiography (DSA) was conducted to identify and assess the ruptured aneurysm. Upon diagnosis, securing the aneurysm to prevent rebleeding was prioritized. Patients were closely monitored in a neurocritical care unit. Blood pressure was tightly controlled to avoid hypertension or significant fluctuations that could increase the risk of rebleeding. Neurological exams were performed regularly, and transcranial Doppler (TCD) was used to detect vasospasm. Additional imaging, including computed tomography perfusion (CTP) and CTA, was utilized as necessary to identify ischemic areas.

### Ultrasound imaging

Ultrasound imaging was performed with a 3D ultrasound scanner (Resolve Stroke, Paris, France) dedicated to transcranial contrast enhanced imaging. A matrix probe with 1024 elements (footprint of 9.6 x 11.1 cm^2^) was driven by the ultrasound scanner at a central frequency of 2.0 MHz. The system allowed a continuous acquisition of 2 mininutes and 45 seconcs at 1000 volume/sec. The non-derated mechanical index was 0.56 at 0.5 cm and 0.2 at 6.5 cm, close to a previous study with 2D ultrasound imaging^34^. The Intensity at Spatial-Peak Time-Averaged (ISPTA) was 241 mW/cm2. The imaging sector represents a pyramidal volume of range 0.5 to 9.0 cm in depth, and a half angle of 20 degrees. During the acquisition, 2.4 ml of ultrasound contrast agent (Sonovue, Bracco, Italy) was injected in a bolus then flush with 10 ml of saline via a peripheral vein catheter. The procedure was repeated similarly on the contralateral side of the head with a pause of a at least 5 minutes between the two recordings to eliminate most microbubbles form the blood stream. For each patient, 3 imaging sessions were performed between days 0-2, days 5-10, and between days 11-15 after the onset.

### Data Analysis

The ultrasound scanner allowed the reconstruction of two display mode from a single set of 3D CEUS data. A Micro Vascular Imaging (MVI) mode is used to highlight the vessels. The MVI is reconstructed from an accumulation of 15s consecutive volumes centered around the bolus peak enhancement. Microbubbles were enhanced by removing the issue signal with a rolling average filter (window size of 11 frames) prior to standard DAS beamforming. The grid size in cartesian coordinate is 9x8x8 *cm*^3^ with an isometric voxel size of 0.5 mm each side. For perfusion analysis, 40 seconds of consecutive volumes were reconstructed around the bolus enhancement peak using the same rolling average filter with a temporal sampling of 256 ms, and an isometric voxel size of 1mm each side. Each frame consists of a power Doppler average over 128 ms. 4D volumes were normalized as follows: (*I − I*_*baseline*_)*/*(*I*_*max*_ *− I*_*baseline*_) , where *I*_*baseline*_ is the averaged signal on 5 frames before the first peak and *I*_*max*_ is the maximum value between the left and right-side acquisitions. This normalization is performed per patient and per time point. To analyze the Time Intensity Evolution (TIE) curves, only enhancements greater than 15% with respect to baseline are considered significant. Outliers are identified on a rolling window as more than 3 times the median value within the window. The start time is defined as the first data point that exceeds 15% above the baseline. The time to maximum is calculated as the difference between the peak time and the start time. The meat transit time is determined by the full width of the curve at 80% of the maximum value.

### Assessment of Ultrasound-Based Prognosis

The Time-Intensity Evolution (TIE) curves derived from Phase 2 3D UPI scans of 11 patients were independently and randomly reviewed by two board-certified intensivists. Each curve was qualitatively rated as Favorable, Unfavorable, or Inconclusive, based on the expected perfusion dynamics. These expert assessments were then compared to the patients’ neurological outcomes, defined using the modified Rankin Scale (mRS) at 6 months, and the Glasgow Outcome Scale – Extended (GOSE) at both 3- and 6-months post-injury.

## Data Availability

All data produced in the present work are contained in the manuscript

## SUPPLEMENTARY MATERIALS

Table 1

**Table 1.** Clinical and demographic characteristics of all patients included in the study.

| Patient | #1 | #2 | #3 | #4 | #5 | #6 | #7 | #8 | #9 | #10 | #11 |
| --- | --- | --- | --- | --- | --- | --- | --- | --- | --- | --- | --- |
| **Delayed Neurological Deficit** | NO | NO | YES | NA | NO | NA | YES | YES | NA | NO | YES |
| **TCD: Vm > 120 cm/s** | YES | NO | NO | NO | NO | YES | NO | YES | YES | NO | YES |
| **TCD: Vm > 200 cm/s<sup>33,35</sup>** | NO | NO | NO | NO | NO | YES | NO | NO | NO | NO | NO |
| **Vasospasm CTA** | NA | NA | NO | YES | NA | YES | YES | NO | NO | NA | YES |
| **Vasospasm DSA** | NA | NA | NA | YES | NA | NA | YES | YES | NA | NA | YES |
| **Original Fisher** | 4 | 4 | 4 | 4 | 4 | 4 | 4 | 4 | 4 | 3 | 4 |
| **MRS Rankin 3m** | 1 | 1 | 1 | 6 | 2 | 6 | 6 | 1 | 6 | 1 | 6 |
| **MRS Rankin 6m** | 1 | 1 | 0 | 6 | 1 | 6 | 6 | 1 | 6 | 0 | 6 |
| **Glasgow Outcome Scale Extended m3** | 6 | 5 | 5 | 1 | 6 | 1 | 1 | 5 | 1 | 5 | 1 |
| **Glasgow Outcome Scale Extended m6** | 7 | 5 | 5 | 1 | 7 | 1 | 1 | 5 | 1 | 5 | 1 |
| **Favorable Neurological Outcome\*** | YES | YES | YES | NO | YES | NO | NO | YES | NO | YES | NO |

\*Favorable neurological outcome was defined as a modified Rankin Scale $\leq 2$ at 6 months.

**Figure Supplementary 1.**
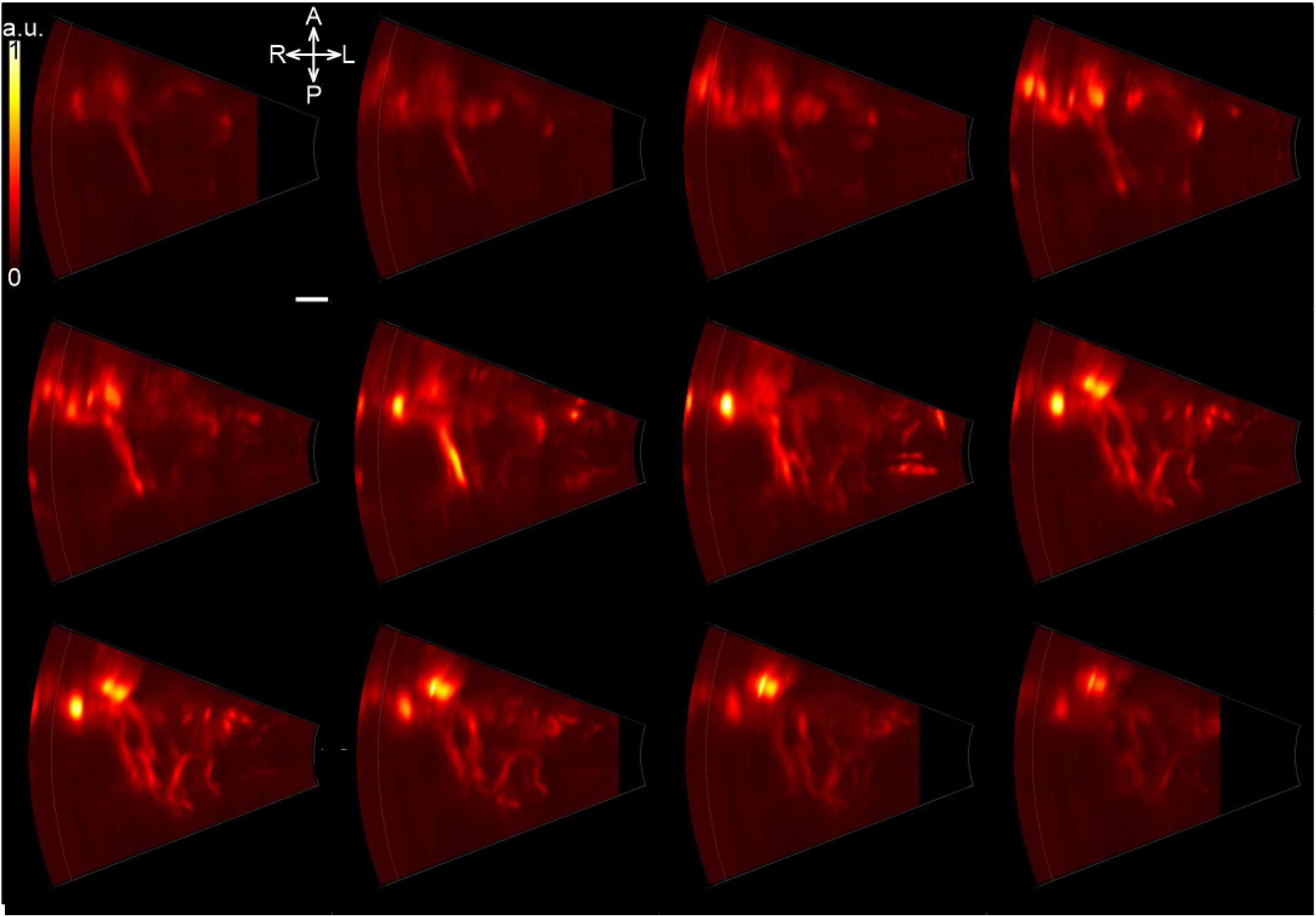
Axial MVI slices of the human brain vasculature acquired through the temporal window. Twelve consecutive axial slices are displayed from superior to inferior, covering a total depth of 39.6 mm with 3.3 mm inter-slice spacing. These Microvessel Imaging (MVI) reconstructions reveal the fine structure of the cerebral vasculature, visualized transcranially through the temporal bone. The sequence captures the vascular organization from superficial cortical regions toward deeper subcortical territories, providing insight into vessel density and distribution across axial planes. The white scale bar correspond to 10 mm.

**Figure Supplementary 2.**
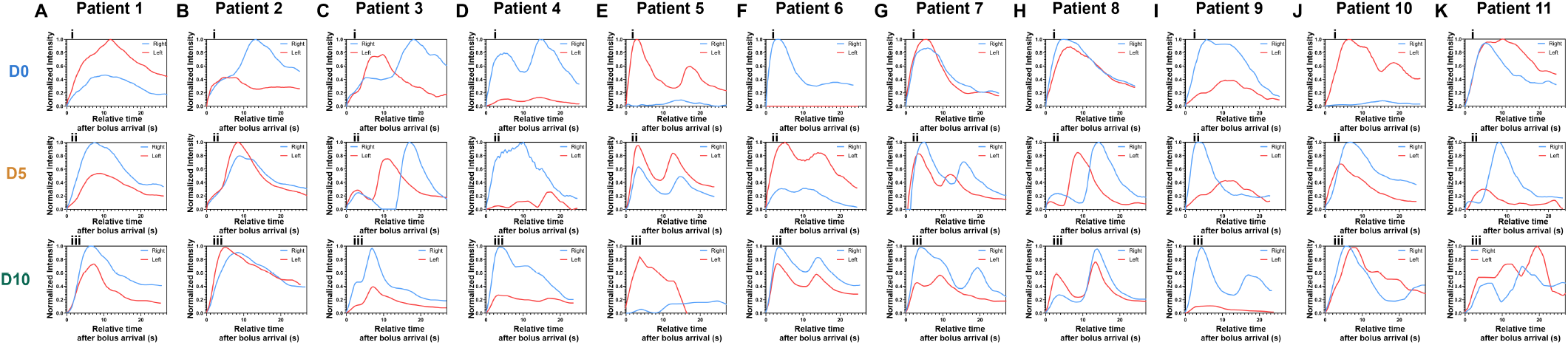
Longitudinal Time-Intensity Evolution curves for all patients (A-K). TIE curves are shown for both hemispheres in all 11 patients across three imaging time points. For each patient, time points i, ii, and iii correspond to the first, second, and third acquisition sessions, respectively. The curves illustrate the evolution of contrast dynamics over time and the degree of inter-hemispheric symmetry or asymmetry observed throughout the follow-up period.

## ACKNOWLEDGMENT

This work was partly supported by the European Research Council (ERC) through the European Union Horizon 586 H2020 Program/ERC Proof-of-concept under Grant 101101066-StrokeMonitor.

